# Understanding Embolus Transport And Source To Destination Mapping Of Thromboemboli In Hemodynamics Driven By Left Ventricular Assist Device

**DOI:** 10.1101/2024.09.23.24314233

**Authors:** Sreeparna Majee, Akshita Sahni, Jay D. Pal, Erin E. McIntyre, Debanjan Mukherjee

## Abstract

Left Ventricular Assist Devices (LVADs) are a key treatment option for patients with advanced heart failure, but they carry a significant risk of thromboembolic complications. While improved LVAD design, and systemic anticoagulation regimen, have helped mitigate thromboembolic risks, ischemic stroke due to adverse thromboembolic events remains a major concern with current LVAD therapies. Improved understanding of embolic events, and embolus movement to the brain, is critical to develop techniques to minimize risks of occlusive embolic events such as a stroke after LVAD implantation. Here, we address this need, and devise a quantitative *in silico* framework to characterize thromboembolus transport and distrbution in hemodynamics driven by an operating LVAD. We conduct systematic numerical experiments to quantify the source-to-destination transport patterns of thromboemboli as a function of: LVAD outflow graft anastomosis, LVAD operating pulse modulation, thromboembolus sizes, and origin locations of emboli. Additionally, we demonstrate how the resulting embolus distribution patterns compare and correlate with descriptors based solely on hemodynamic patterns such as helicity, vorticity, and wall shear stress. Using the concepts of size-dependent embolus-hemodynamics interactions, and two jet flow model for hemodynamics under LVAD operation as established in our prior works, we gain valuable insights on departure of thromboembolus distribution from flow distribution, and establish that our *in silico* model can generate deep insights into embolus dynamics which is not otherwise available from standard of care imaging and clinical data.

## 1 Introduction

Heart failure constitutes a major global health concern, affecting at least 26 million individuals worldwide [1–3]. The prevalence of heart failure continues to rise over time with the aging population. According to latest statistics compiled by the American Heart Association (AHA) and the National Health and Nutrition Examination Survey (NHANES), an estimated 6.7 million American adults above the age of 20 years (2.2%) had advanced heart failure between 2017 and 2020 [4]. Left ventricular assist devices (LVADs) are implantable pumps that provide mechanical circulatory support and have emerged as the standard of care for bridge-to-transplant as well as destination therapy in advanced heart failure patients [5–8]. Although there have been significant advancements in LVAD therapy over the past several decades, ischemic stroke remains a significant cause of morbidity and mortality in patients after LVAD implantation [9, 10]. Stroke caused after LVAD implantation have been reported in multiple studies with a rate in the range of 11-47% [11–14]. The most recent report of the Interagency Registry of Mechanical Circulatory Support (INTERMACS), which included the least thrombogenic LVADs currently available (HeartMate3) the stroke rate was still noted at 6-8% [15]. Progress in understanding and reducing the risks of post-implant strokes can have a profound effect on the overall success of treatments. However, a significant obstacle lies in the limited understanding of the factors that contribute to post-implant stroke risks in individuals supported by LVAD. For increased surgical treatment efficacy, understanding and considering these factors prior to the surgical implantation is advantageous.

Thrombotic complications in LVADs can be attributed to various factors, including the activation of the extrinsic coagulation pathway due to foreign materials used in the pump (*hemocompatibility*), shear stress, flow stasis in the ventricle due to ventricular dysfunction, stasis in the pump body, or the outflow graft [16]. Concurrent inflammation and infection can also promote coagulation [17, 18]. Shear-induced hemolysis leads to the release of ADP from red blood cells, subsequently triggering platelet activation [17]. Additionally, altered flow patterns resulting from LVAD operation can contribute to endothelial injury within blood vessels, potentially accelerating the development of thrombus in the LVAD graft [19, 20]. Consequently, the LVAD outflow graft represents a major source of thromboemboli that can lead to cerebrovascular accidents and embolic strokes. Furthermore, flow stasis observed in the aortic arch root due to altered hemodynamics [21] after LVAD implantation can lead to thrombosis in the aorta. Intermittent aortic valve opening from ventricular contractions can then propel thromboembolic particles in the aortic root towards the cervical vessels. Improved understanding of stroke risks in patients in LVAD support, therefore, will require understanding how thromboemboli originating from these different embolization sites can distribute towards the brain.

Such insights on stroke post-LVAD will require quantitative characterization of post-implant hemodynamics, as well as thromboembolic events. Thrombotic events, risks of embolization, and embolus distribution to the brain, are not just a function of patient vascular anatomy and physiological variables, but also intimately depends upon hemodynamics and embolus-hemodynamics interactions. However, standard imaging and clinical workup, as well as *in vivo* investigations using animal models do not always provide these information in sufficient resolution. *In silico* advances have enabled a viable avenue for studying such phenomena in a patient-specific manner. Extensive *in silico* investigations have been reported on investigating the impact of the surgical attachment angle of the LVAD outflow graft on hemodynamics within the aortic arch and its potential for thromboembolic events [22–27]. Computational research has delved into various specific factors, including but not limited to: examining the impact of pulse modulation on LVAD flow in both adult and pediatric populations [28–30], exploring additional surgical attachment parameters for VADs [31], investigating the influence of viscoelastic hemorheology on VAD-powered circulation [32], and assessing the consequences of intermittent reopening of the aortic valve [33]. In a series of prior works, we have developed a computational framework to quantitatively assess a range of hemodynamic descriptors such as helicity, vorticity, wall shear, and viscous energy dissipation as function of LVAD outflow graft anastomosis and pulse flow modulation [21, 34]. However, detailed *in silico* quantification of spatiotemporally varying embolus-hemodynamics interactions and thromboembolus transport towards the cervical vessels remains sparingly investigated in LVAD driven circulation. Motivated by this knowledge gap, here we demonstrate a patient-specific *in silico* embolus-hemodynamics model, established extensively in our prior works for stroke [35–38], for quantitative characterization of embolus distribution towards the cervical vessels post-LVAD implantation. Our goal is to demonstrate key features of thromboembolus source to destination transport trends as function of surgical variables such as varying graft anastomosis and pulse flow modulation, and embolus properties such as size and release locations.

## 2 Methods

### 2.1 Image-based modeling of vascular anatomy

For this study, we selected a patient-specific vascular network used extensively in our prior works for quantitative assessment of hemodynamics driven by LVADs [21, 34], adapted from the open source SimVascular model repository [39]. The native vascular anatomy for the model considered here includes the aortic arch and branch arteries extending up to the iliofemoral arteries. Computed Tomography (CT) images of the selected vascular network were segmented using a 2D lofted image segmentation technique, which were subsequently used to generate a patient-specific 3D model using the open source software tool SimVascular [40]. An LVAD outflow graft was then surgically placed along the aortic arch of the patient-specific model, using an image-based virtual surgical anastomosis workflow as detailed in our prior study [21]. The LVAD outflow graft was virtually attached such that the resulting graft does not intersect nearby organ (*that is, heart, lungs etc.*) or bone (*that is, sternum, ribs etc.*) boundaries. This ensured generating realistically achievable surgical LVAD outflow graft anastomoses. Using this approach, we generated a set of 9 different parametric graft anastomoses, based on 3 combinations of orientation angles measured towards or away from the aortic valve (*labelled with an Inc tag*), and 3 combinations of orientation angles measured towards left or right of the heart across the coronal plane (*labelled with an Azi tag*). The model generation, and the varying anastomosis angles, are illustrated in Fig. 1 (a) and (b). Specifically, the graft anastomosis angles for inclination angles are as follows: (1) perpendicular to the aorta (*Inc90*); (2) 45*^○^* towards the aortic valve (*Inc45*); and (3) 45*^○^* towards the aortic arch (*Inc135*). The graft anastomosis angles for azimuthal angles are as follows: (1) 45*^○^* right of the heart (*AziNeg45*); (2) perpendicular to the coronal plane (*Azi0*); and (3) 45*^○^* left of the heart (*Azi45*). Furthermore, in this study, we assumed that the aortic valve was sealed throughout and has no aortic valve opening. This assumption, as well as additional intricacies of the image-based workflow, has been discussed at length in our prior works [21, 34].

**Figure 1:**
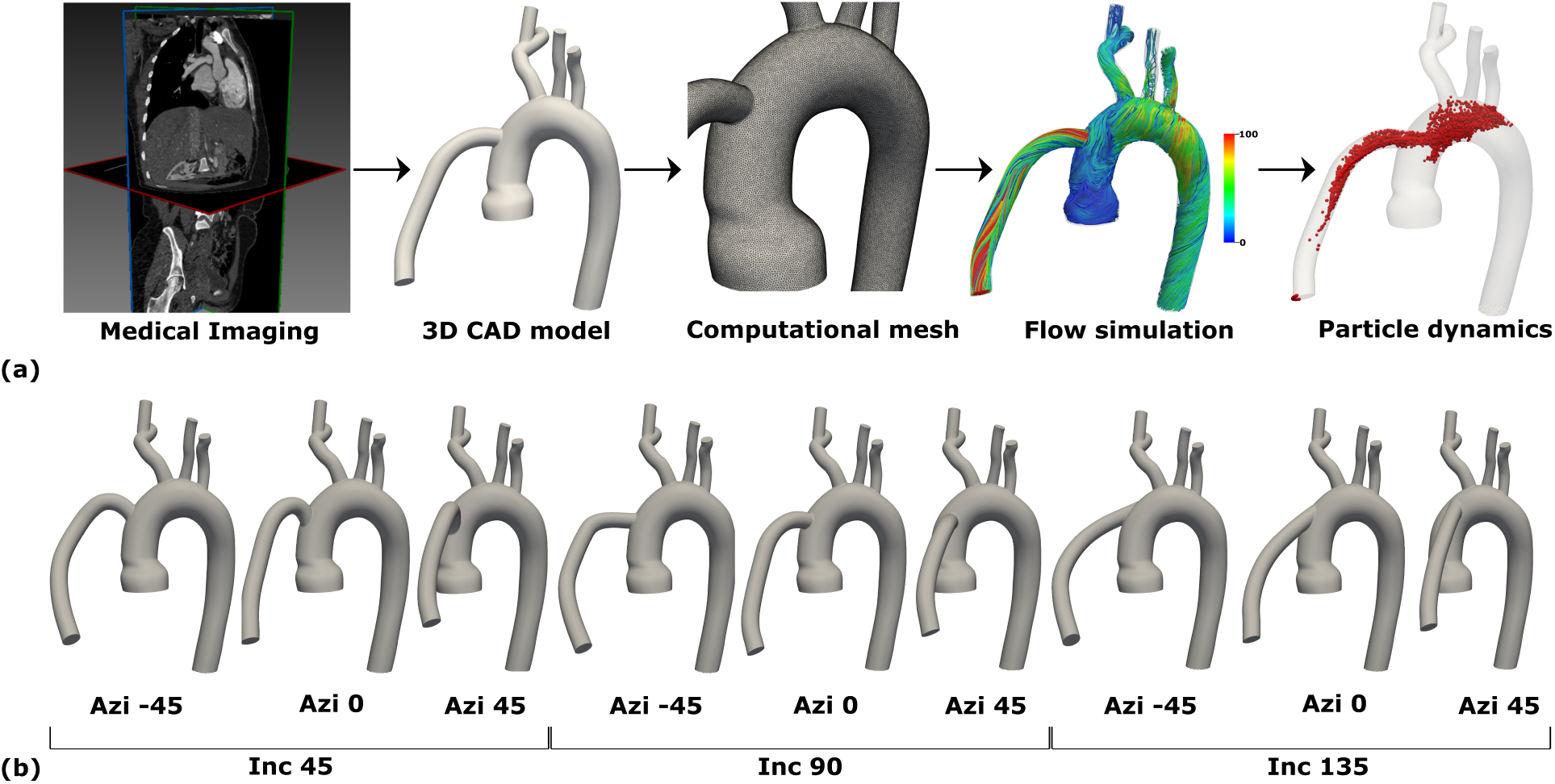
(a) Schematic overview of the overall computational framework, (b) A virtual family of LVAD models with varying graft anastomosis.

### 2.2 Patient-specific hemodynamics modeling

The overall workflow starting from image data to embolus transport model has been illustrated in Figure 1(a). Three-dimensional simulations of blood flow was conducted for each of the 9 models, assuming blood to be a homogeneous Newtonian fluid [41] of effective density (*ρ*) 1060 kg/m^3^ and dynamic viscosity (*µ*) 4.0 cP. The flow domain comprising the the vessel lumen and the LVAD outflow graft for each of the 9 models were discretized into a computational grid comprising linear tetrahedral elements with a maximum edge size of ≈ 0.67 mm. For each model, blood flow velocity and pressure was computed by solving the incompressible Navier-Stokes equations for momentum balance and the continuity equation for mass balance. A Petrov-Galerkin stabilized finite element method [42–45] implemented in the open source SimVascular package [40], was used to solve these equations, using a numerical integration time-step of 0.001 sec. Flow was driven from the LVAD outflow graft inlet mapped spatially into a Womersley-type flow profile. As described in detail in our prior works [21, 34], a set of three different scenarios of pulse modulation were considered for the inlet flow from the LVAD outflow graft: (a) a constant uniform flow over time; (b) flow with a low extent of pulse modulation; and (c) flow with a high extent of pulse modulation; with the pulse modulated flow profiles adapted from prior studies [46]. The time-averaged inlet flow was fixed at 4.9 L/min for all the models. Three-element Windkessel boundary conditions with parameters: proximal resistance (*R_p_*); distal resistance (*R_d_*); and compliance (*C*); were used to consider the effect of the truncated downstream vasculature on the flow. Quantitative details of the three inflow profiles with their pulsatility index (PI), and the individual Windkessel parameters *R_P_*, *R_d_*, and *C* have been specified in our prior work [21, 34] and are not reproduced here for brevity. The numerical simulation was implemented using the built-in stabilized finite element solver in SimVascular. Blood flow was simulated for a total of 9 different anastomosis models, with 3 different inflow patterns, leading to 27 different CFD models, each simulated for a duration of 3 seconds. The results from the third second of the total simulation duration was further used for embolus transport calculations outlined next.

### 2.3 Modeling the movement of thromboemboli

Transport of thromboemboli across the arteries was modelled, by assuming each embolus to be a spherical particle, and modeling their dynamics in the simulated blood flow using a custom modified form of the Maxey-Riley equation [47], that we have developed extensively in a series of prior works [35–38]. Briefly, this equation incorporates the effects of the drag force, shear-gradient-driven lift forces, fluid stresses for undisturbed flow, added mass and buoyancy forces, alongwith a soft particle-wall collision model to account for embolus interactions with vessel wall. The final form of this equation is stated as follows:

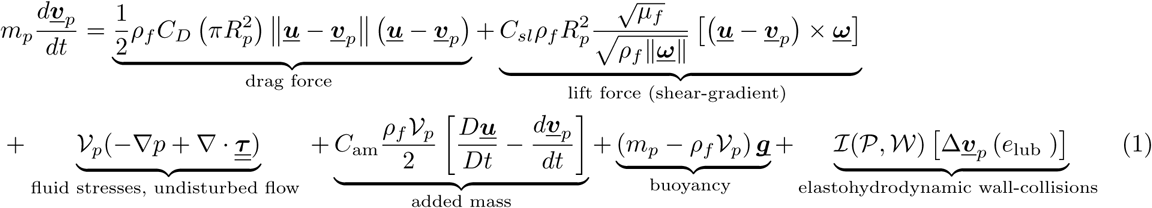

In the above equation, *m_p_* denotes the particle mass, *R_p_* is the particle radius, ν*_p_* is the particle volume (*which in this case will be* 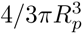 *with the spherical shape assumption*), *τ* denotes the stress tensor from undisturbed forces (*obtained from CFD simulations*), ***<u>u</u>*** is the simulated blood flow velocity output from CFD simulations, and **v***_p_* is the translational particle velocity. C_*am*_ is the added mass coefficient (which has a fixed value of 0.5 for spherical particle shape), *C_D_* is the drag coefficient, and *C_sl_* denotes the shear-gradient lift force coefficient adopted from prior works [48, 49]. ***<u>ω</u>*** = Δ × ***<u>u</u>*** is the vortcity of the flow where the particle is located. *I*(*P*, *W*) is an indicator that has a value of 1 when a particle *P* comes into contact with the artery wall, and the value of this function is 0 otherwise. Next, the contribution of the momentum changed created by the elastohydrodynamic lubrication and particle-wall collisions is denoted by the term [δ**v***_p_*(e_lub_)] where e_lub_ is the restitution coefficient. For the embolus transport modeling in LVAD driven flows, precise resolution of the embolus interactions with the vessel walls is especially critical in the regions at and around the LVAD outflow jet impingement on the aorta wall. To address this, we modified the particle-wall contact resolution method from our prior works [36] into one that relies on computed Signed Distance Fields (SDF) of the vascular geometry. Specifically, we use a fast voxel-based interpolation method [50] to compute the SDF based on the 3D model created from image-segmentation; interpolate the computed SDF back on to the CFD grid over which flow and particle dynamics computations occur; and resolve particle-wall contact by interpolating the computed SDF at particle locations and checking if computed SDF is less than particle radius *R_p_*. Each thromboembolus particle is released individually from a specified source or release location, making this a Monte Carlo type sampling based simulation; and hence, no contact mechanics interactions between individual thromboemboli were considered.

### 2.4 Design of in-silico experiments

Using the underlying image-based modeling, hempodynamics simulation, and embolus transport simulation techniques as identified in Sections 2.1, 2.2, and 2.3 respectively; we setup a parametric design for *in silico* experiments to understand embolus dynamics patterns in LVAD driven flows. Flow from the LVAD outflow graft leads to an altered state of hemodynamics when compared against normal physiological flow, which we have conceptually substantiated using a *Two Jet Flow Model* for LVAD hemodynamics in our prior works [21, 34]. Specifically, we note that when compared against flow driven by the opening of the aortic valve in normal scenario (*the Aortic Outflow (AO) jet*); hemodynamics driven by the LVAD outflow (LO) jet leads to significantly differing flow patterns owing to the jet traversing across the aorta centerline, impinging with the aorta wall, and subsequent flow roll-up. Consequently, this can lead to regions of stasis in the aortic root, especially for cases where the outflow graft angles away from the aortic valve, leading to the impingement site moving away from the aortic root (*indicated in detail in parametric analysis showed in* [21]). This motivates our *in silico* study design, by accounting for these hemodynamic features and their connection to thrombembolic events explicitly in the source-destination mapping analysis. Specifically, we consider the following scenarios. First: as flow enters the aorta from the LVAD, thromboemboli originating from device-hemodynamics interactions can enter the flow direcctly via the LVAD outflow graft inlet. Some of these emboli can travel directly towards the brain via cervical vessels, while some may reach the aortic root region and remain accumulated there. Second: thrombogenicity induced by flow stasis in the aortic root can lead to additional thromboemboli in the aortic root. Third: the accumulated emboli from the *first* and *second* factors above can get re-entrained in blood flow in case of intermittent aortic valve opening, and subsequently reach the brain via the cervical vessels. While under high LVAD support, the valve remains mostly closed (*which is what we have assumed for this study*), this risk of embolic events need to be nevertheless included in the analysis. Motivated by these, we consider two thromboembolus sources: LVAD outflow graft inlet, and the aortic root region; and two destinations of relevance for stroke: the branching arteries into the cervical vessels, and the aortic root region. For each source-destination combination, we conduct thromboembolus transport for each of the 9 different anastomosis, for each of the 3 pulse modulation scenario. We consider a set of 8 different thromboembolus sizes ranging from 0.5 mm to 4.0 mm in diameter, mimicking a range of stroke scenarios. The outline of this parametric design of experiments is presented in Figure 2. Across all parameter combinations, this led to a total of 432 simulations, for each of which 5,000 thromboembolus samples were tracked, leading to source to destination distribution data spanning a total of 2.16 million embolus samples. Each thromboembolus was modeled as idealized sphers of material density 1.1 g/cm^3^ as obtained from literature [51, 52]. For each of the 27 models, flow velocity data from the third second of simulated hemodynamics was stitched together 4 times to create a representative flow-field spanning 4 cardiac cycles that has no cycle-to-cycle flow variations. For each of these 432 simulations, the computational endpoint comprised a number fraction defined as *N_k_*/*N_res_* where *N_k_* is the number of emboli that reached the cervical vessels or accumulated in the aortic root from the LVAD graft or aortic root source and *N_res_* is the total number of emboli that is resolved in each variation of the 3D LVAD model after 4 cardiac cycles. Details of these calculations are also provided in our prior studies [53].

**Figure 2:**
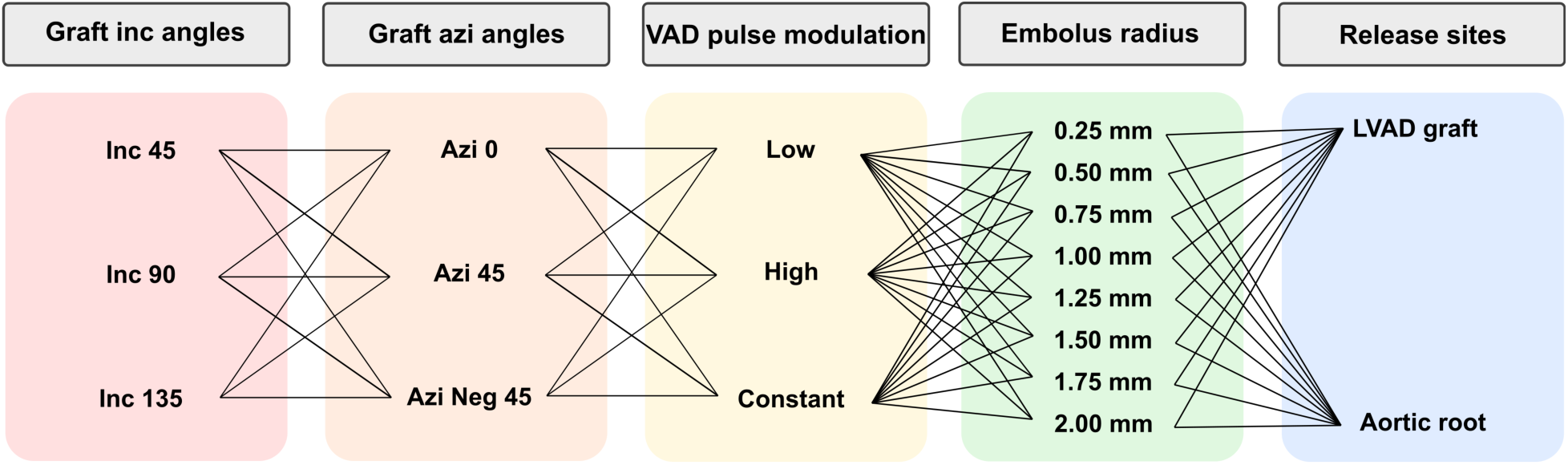
The structure of the in silico experiments comprising a total of 432 different embolus transport simulations. The parameters include three inc angles, three azi angles, three LVAD pulse modulation, eight embolus sizes, and two release locations.

## 3 Results

### 3.1 Thromboembolus distribution from LVAD outflow graft

Here we discuss simulated thromboembolus distribution from the LVAD outflow graft as the source, as function of varying outflow graft anastomosis and pulse modulation of the LVAD. Figure 3(a) depicts the distribution of thromboemboli from outflow graft moving into the cervical vessels towards the brain across all 27 flow scenarios, combining all thromboembolus particle sizes. The corresponding thromboembolus distribution from LVAD outflow graft towards the aortic root is presented in Figure 3(b). Based on these simulated data, we note that distribution of thromboemboli towards the cervical vessels is significantly greater than that of accumulation in the aortic root, when emboli entering from the LVAD outflow graft are considered. The data also indicate that both the anastomosis angles, as well as the pulse modulation, influences the distribution of thromboemboli coming in through the outflow graft. Across the *Inc* anastomosis angles considered, for all combinations and across all particle sizes, the *Inc135* case - that is, angled away from the aortic valve at 45*^○^* towards the aortic arch - has least extent of emboli distributing towards the cervical vessels. The combination with the highest extent of thromboembolus distribution towards the cervical vessels is the *Inc45AziNeg45* anastomosis with low pulse modulation; while that with the least extent of distribution towards the cervical vessels is *Inc135Azi-45* anastomosis with low pulse modulation. While accumulation in the aortic root is significantly lesser, it is relevant to note that *Inc135* anastomoses with graft angled away from the aortic valve leads to higher extent of accumulation compared to other anastomoses, across all pulse modulation and particle sizes considered.

**Figure 3:**
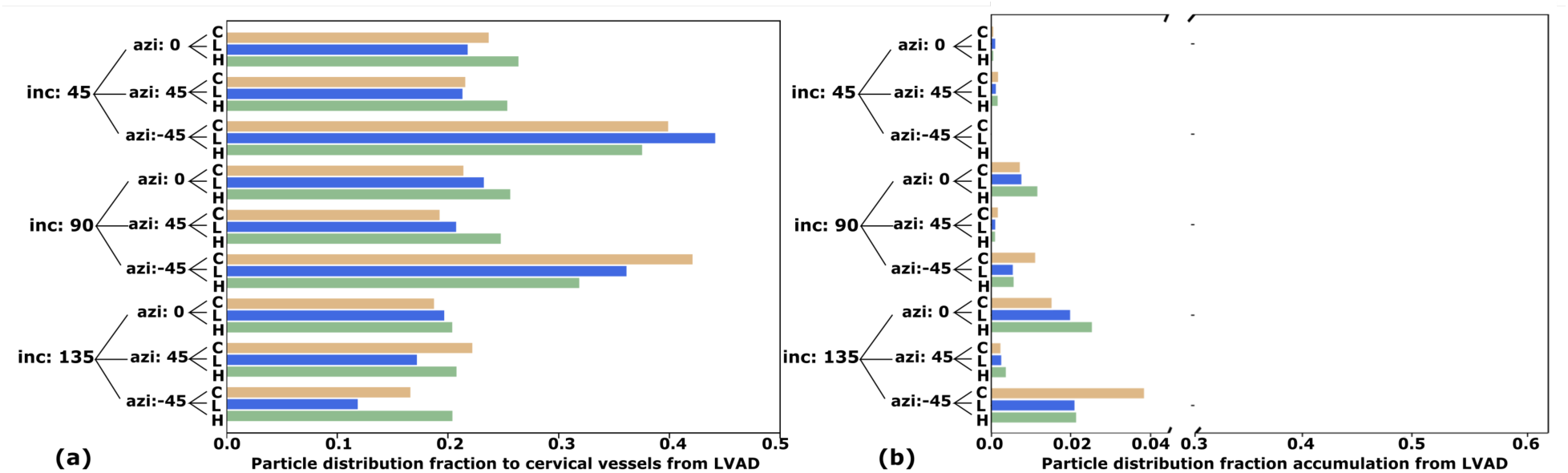
(a) Total distribution of emboli to cervical vessels across all particle size originating from LVAD graft; (b) Total accumulation distribution of emboli in aortic root across all particle size originating from LVAD graft. The distribution is represented as fraction of emboli that reached the cervical vessels to the total amount of resolved particles at the end of 4 cardiac cycles.

### 3.2 Thromboembolus distribution from aortic root

Similar to Section 3.2, the simulated thromboembolus distribution from the aortic root region as the source is presented in Figure 4, illustrated as function of varying outflow graft anastomosis and pulse modulation of the LVAD. Similar to the trends observed in Section 3.1, we note that the distribution of thromboemboli from aortic root region as the source is influenced by both anastomosis angles, as well as extent of pulse modulation. The distribution of thromboemboli into the cervical vessels towards the brain is presented in Figure 4(a), while the acccumulation of thromboemboli in the root region (those that remain entrained at the source) is presented in Figure 4(b). We note that thromboembolus accumulation at the root is significantly higher for emboli originating at the root, compared to those entering from the LVAD outflow graft. The extent of accumulation is also markedly higher for the *Inc135* anastomoses, with the LVAD outflow graft angled away from the aortic root. Conversely, for these *Inc135* anastomoses, the extent of distribution towards the cervical vessels is notably lower, with the least extent of thrombembolus distribution towards the cervical vessels noted for the *Inc135Azi0* anastomosis with low pulse modulation. The *Inc45Azi-45* and the *Inc90Azi-45* anastomoses lead to the highest extents of thromboemboli moving potentially towards the brain, with the low pulse modulation scenario for both cases having a distribution fraction of 0.47 - the maximum distribution fraction towards the cervical vessels from the aortic root. Likewise, the case with highest embolus accumulation is the *Inc135Azi0* low pulse modulation scenario with a computed fraction of 0.62 across all thromboembolus sizes.

**Figure 4:**
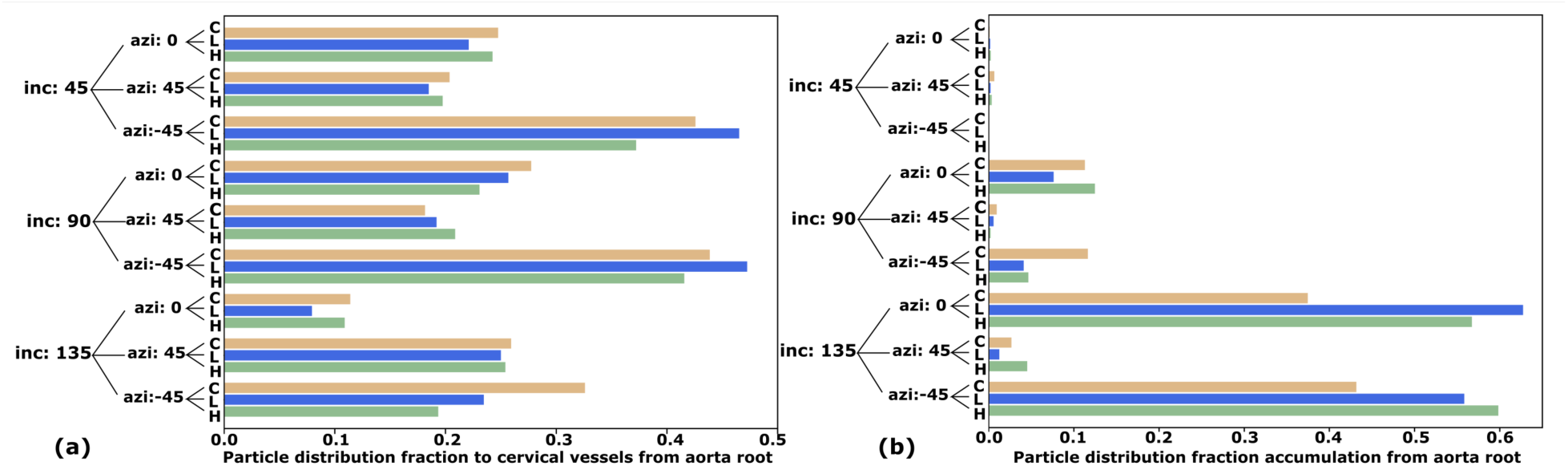
(a) Total distribution of emboli to cervical vessels across all particle size originating from Aortic root; (b) Total accumulation distribution of emboli in aortic root across all particle size originating from Aortic root. The distribution is represented as fraction of emboli that reached the cervical vessels to the total amount of resolved particles at the end of 4 cardiac cycles.

### 3.3 Combined thromboembolus distribution governing stroke risk

Here we illustrate the combined source to destination distribution for both sources (*LVAD outflow graft and aortic root*) and across both thromboembolus destinations (*cervical vessels and aortic root*). These are computed as the fraction of emboli that reach the cervical vessels or the aortic root from both sources considered in our simulations. The combined distribution is illustrated in Figure 5 as function of varying out-flow graft anastomosis and pulse modulation of the LVAD. Figure 5 (a) presents combined thromboembolus distribution towards the cervical vessels, across all of the 27 flow scenarios considered here and combining all thromboembolus particle sizes. The corresponding combined accumulation in the aortic root is presented in Figure 5 (b). We observe that outflow graft anastomoses with angle *Azi-45* have greater extent of embolus distribution to the cervical vessels from both sources, across all thromboembolus sizes combined. Additionally, while the anastomosis angled 45*^○^* towards the aortic arch (*that is Inc135 models*) altogether have least extent of thromboembolus distribution directly towards the arch, they have the greatest extent of thromboembolus accumulation at the aortic root. This indicates that expectations based on anastomosis angles alone can lead to competing trends in embolus transport patterns when direct traversal towards the cervical vessels and accumulation in the aortic root region are simultaneously considered. The maximum distribution of thromboemboli form both sources combined, towards the cervical vessels, is observed for the *Inc45AziNeg45* model with low pulse modulation. The maximum accumulation of thromboemboli from both sources at the aortic root is observed for the *Inc135Azi0* model with low pulse modulation.

**Figure 5:**
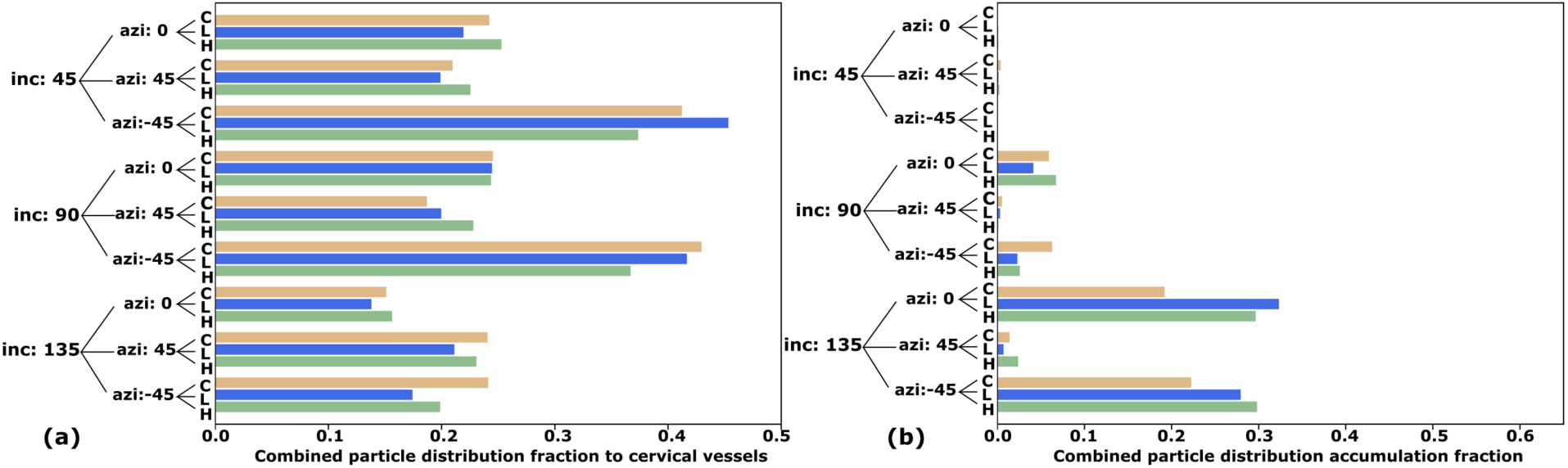
(a) Total distribution of emboli to cervical vessels across all particle size originating from both LVAD graft and aortic root combined; (b) Total accumulation distribution of emboli in aortic root across all particle size originating from both LVAD graft and aortic root combined. The distribution is represented as fraction of emboli that reached the cervical vessels to the total amount of resolved particles at the end of 4 cardiac cycles.

### 3.4 Effect of pulse modulation on thromboembolus fate

As outlined in 2.2, three different LVAD pulse modulations were considered for the simulations, to quantify differences in thromboembolus transport to the cervical vessels due to extent of pulse modulation. The results presented in Figures 3, 4, and 5 together indicate that varying extent of pulse modulation, for the different anastomoses considered, lead to significant variabilities in embolus source to destination transport. Specifically, for thromboembolus distribution from LVAD outflow graft to the cervical vessels, high pulse modulation was associated with higher distribution fractions (*compared to constant and low pulse modulation*) for 6 out of the 9 anastomoses considered. Likewise, for thromboembolus distribution from aortic root to the cervical vessels, constant (*no pulse modulation*) was associated with higher distribution fractions (*compared to high and low modulation*) for 6 out of the 9 anastomoses considered. In both of these cases, the highest distribution of thromboemboli to the cervical vessels were associated with low pulse modulation cases - *Inc45AziNeg45* for thromboemboli from LVAD outflow graft, and *Inc90AziNeg45* for thromboemboli from aortic root. While these observations indicate that pulse modulation influences extent of embolus movement towards the brain in LVAD flows, there are no consistent trends or dependency on the extent of pulse modulation that can be inferred.

### 3.5 Effect of thromboembolus size on thromboembolus distribution

The simulated thromboembolus distribution shows a strong dependence on embolus size, which is in agreement with observations on embolus transport in arterial flows as indicated in several prior works [35, 38, 54]. Figure 6 illustrates the variation of thromboembolus distribution towards the cervical vessels from LVAD outflow graft, aortic root, and both sources combined. Figure 7 illustrates the same for thromboembolus accumulation at the aortic root. The trends are presented as function of the outflow graft angle towards/away from the aortic valve. The distribution shows a strong size-dependence for both movement into the cervical vessels, and accumulation at the aortic root. When thromboembolus distribution into the cervical vessels is considered, the average trend across all anastomoses and pulse modulation combinations indicates a decrease in distribution with increasing embolus size. When individual anastomoses are considered, we observe that for *Inc45* and *Inc90* cases, the thromboembolus distribution with size departs from this average trend, and for the size ranges considered here, thromboembolus movement into the cervical vessels increases with size - for both LVAD outflow graft, aortic root, as well as both sources combined. Conversely, thromboembolus distribution to the cervical vessels for all *Inc135* configurations consistently reduce with increasing size. On the other hand, considering thromboembolus accumulation at the aortic root illustrated in Figure 7, we observe that the average trend across all anastomoses and pulse modulation combinations indicates an increase in accumulation with increasing embolus size. Cases with *Inc135* anastomoses, as noted in Section 3.1 and 3.2, have low extents of accumulation at the root overall.

**Figure 6:**
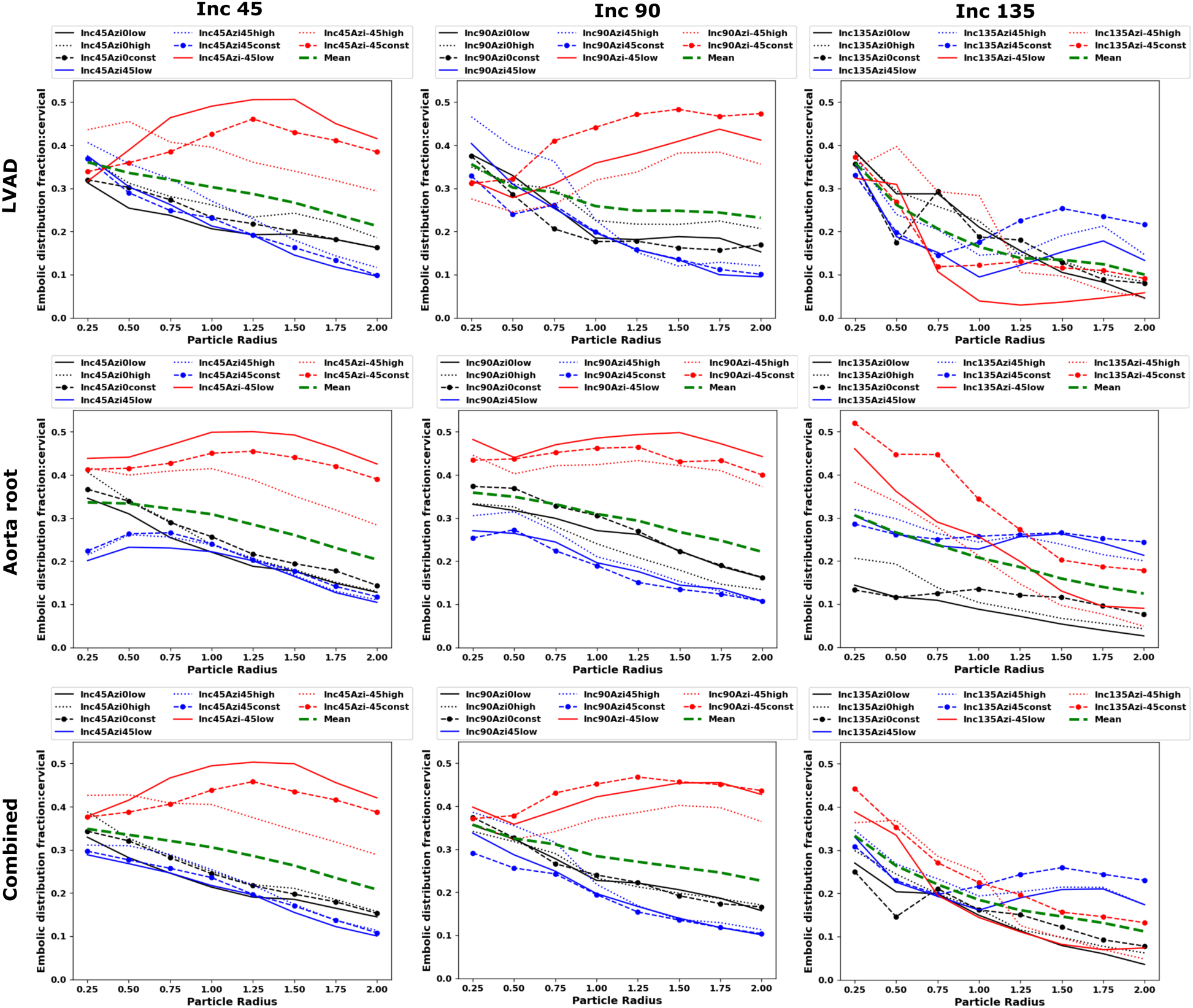
Distribution of embolus particles along with embolus size based on inclination angles for LVAD source, aortic root source and combined source towards cervical vessels. The color black, blue and red represents three different Azi angles: Azi0, Azi45 and Azi-45 respectively. The solid line, dashed line and line with marker represents low pulse modulation, high pulse modulation and constant flow respectively. The mean of the plots in each panel is represented in dashed line with color green.

**Figure 7:**
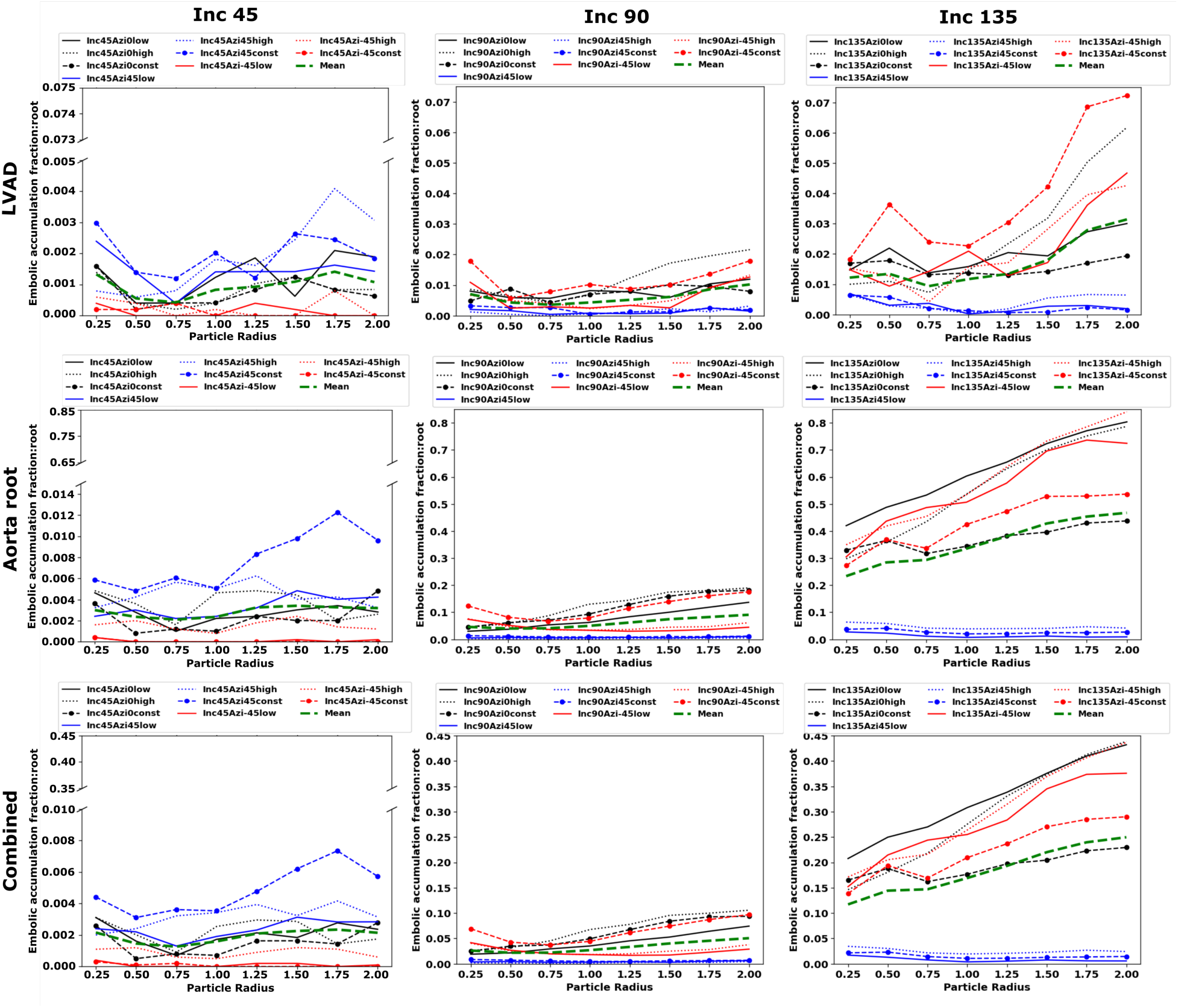
Accumulation of embolus particles along with embolus size based on inclination angles for LVAD source, aortic root source and combined source in aortic root. The color black, blue and red represents three different Azi angles: Azi0, Azi45 and Azi-45 respectively. The solid line, dashed line and line with marker represents low pulse modulation, high pulse modulation and constant flow respectively. The mean of the plots in each panel is represented in dashed line with color green.

**Figure 8:**
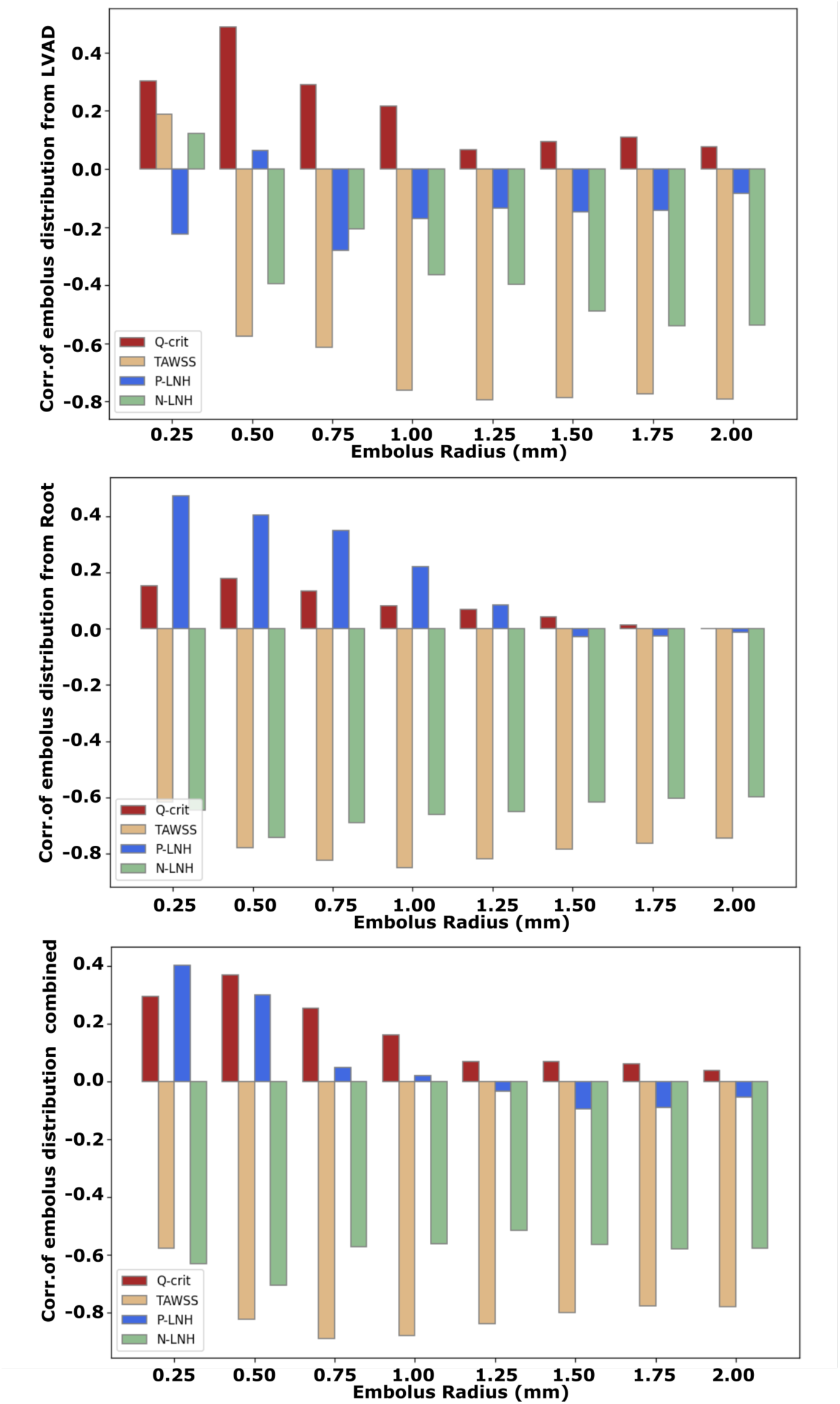
Correlation of embolus distribution fraction towards cervical vessels with ratio of hemodynamic descriptors: Q-criteria, Time averaged wall shear stress, positive local normalized helicity and negative local normalized helicity with varying embolus size.

### 3.6 Correlation of embolus distribution with hemodynamic alteration

In a prior work [21], we have focused exclusively on hemodynamic descriptors for varying outflow graft angles and pulse modulation. With the extensive *in silico* analysis of thromboembolus distribution as conducted here, we can now further analyze the correlation between embolus distribution and hemodynamic descriptors in LVAD driven flows. We compute correlation coefficients between the computed thromboembolus distribution and the hemodynamic descriptors as defined in our prior work [21]. These quantified extent of vorticity using Q-criteria, positive and negative helicity using LNH, and wall shear stress using TAWSS, each compared between LVAD-driven flow and baseline flow without LVAD. The computed Pearson linear correlation coefficients between these descriptors and source-to-destination thromboembolus distribution is listed out in Table 1. We note that, based on the simulation data, thromboembolus distribution shows weak to no correlation with vorticity descriptor; while showing some of the strongest correlation with wall shear stress descriptor. Thromboembolus accumulation at the aortic root shows moderate correlation with positive helicity, while negative helicity descriptors are more correlated with thromboembolus distribution to the cervical vessels. We further note that these correlations also present with strong thromboembolus size dependency, which is illustrated in Figure 9. This illustration affirms the strong negative correlation with wall shear descriptors, and overall negative levels of correlation with negative helical flow, with variations in thromboembolus size.

**Figure 9:**
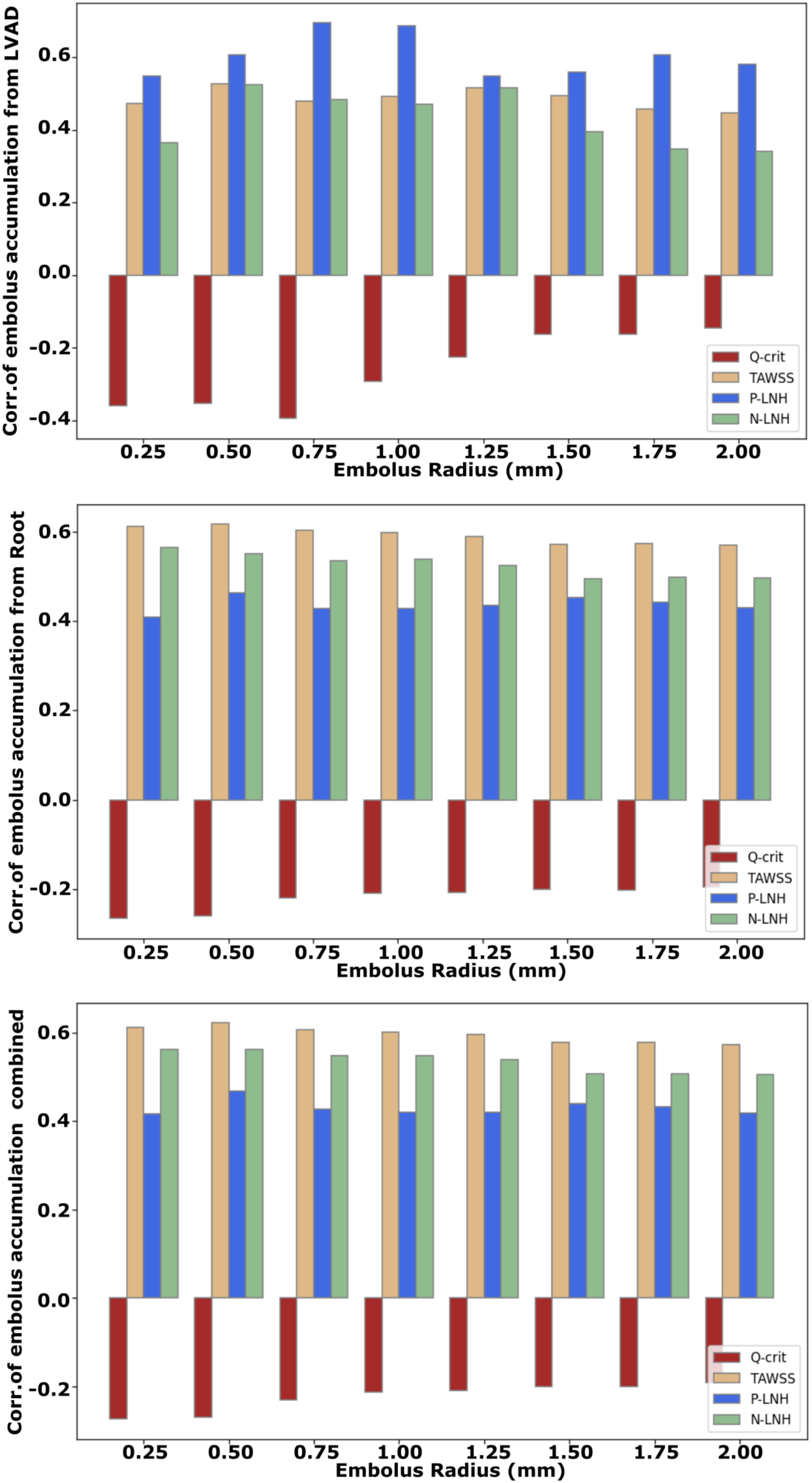
Correlation of embolus accumulation fraction in aortic root with ratio of hemodynamic descriptors: Q-criteria, Time averaged wall shear stress, positive local normalized helicity and negative local normalized helicity with varying embolus size.

**Table 1:** Correlation of source to destination distribution of thromboemboli with hemodynamic descriptors for extent of vorticity (ratio of Q-criteria between LVAD driven flow and baseline flow without LVAD), extent of wall shear stress (ratio of TAWSS between LVAD driven flow and baseline flow without LVAD), extent of positive helicity and negative helicity (ratio of +ve/-ve local normalized helicity between LVAD driven flow and baseline flow without LVAD). Thromboembolus distribution is represented here combined across all embolus sizes considered. All correlation estimated using Pearson’s linear correlation coefficients.

| Destination | Source | Vorticity corr. | TAWSS corr. | +ve helicity corr | -ve helicity corr. |
| --- | --- | --- | --- | --- | --- |
| Cervical | LVAD | 0.20 | -0.83 | -0.17 | -0.48 |
| Cervical | Root | 0.08 | -0.82 | 0.17 | -0.69 |
| Cervical | Combined | 0.14 | -0.87 | 0.02 | -0.63 |
| Root | LVAD | -0.24 | 0.50 | 0.63 | 0.43 |
| Root | Root | -0.21 | 0.59 | 0.44 | 0.52 |
| Root | Combined | -0.22 | 0.60 | 0.43 | 0.53 |

## 4 Discussion

### 4.1 Insights on hemodynamics and the two jet flow model

We present thromboembolus transport simulations in a patient-specific vascular network comprising the aortic arch and branching arteries with a virtually attached LVAD outflow graft. The results shed several insights into embolus transport and source to destination distribution patterns when the flow is driven by an operating LVAD for varying extent of pulse modulation, and varying thromboembolus sizes. As noted in several of our prior works [36–38], the transport of emboli from a source to a destination site is intimately related to the overall state of hemodynamics in the vessels that the emboli travel through. For circulation driven by an operating LVAD, blood flow patterns in the aorta and branching vessels are determined by the impingement of a jet flow emanating from the LVAD outflow graft on the aorta wall. In our previous contributions [21, 34] we have described a *“two-jet flow model”* for LVAD hemodynamics, to explain how flow under LVAD operation driven by this outflow graft jet differs from baseline physiological flow (*in the absence of an operating LVAD*) driven by jet flow emanating from the aortic valve opening. The outflow graft anastomosis angle determines the angle and location of the outflow jet entry and its impingement on the aorta wall. Extent of pulse modulation will influence the temporal intensity of the impingement flow. Subsequently, as a thromboembolus enters the aortic arch through the LVAD graft, it can get entrained in the flow patterns of the aortic arch and ultimately travel either to the cervical vessels, aortic downstream or get accumulated in the aortic root region due to flow stasis. These factors explain the strong dependence of embolus source to destination distribution on both anastomosis angles and pulse modulation as observed in our study. Additionally, we have leveraged the assumption that the aortic valve remains closed for all simulation cases considered here, based on evidence on aortic valve closure under high LVAD support [55, 56]. Hence, in our simulations only LVAD outflow jet flow is considered, with no concurrent aortic valve jet flow. However, the potential effect of intermittent valve opening, and presence of an aortic jet that can re-entrain accumulated thromboemboli at the aortic root has been accounted for in our simulated data by treating the aortic root itself as a potential source or site of thromboembolization. These interpretations enable us to connect our embolus transport observations to the previously outlined two jet LVAD flow model.

### 4.2 Insights on embolus-hemodynamics interactions

Finite sized embolus particles interact with local blood flow features as a function of their size and inertia, and this embolus-hemodynamics interaction mechanics determines embolus fate. We have characterized the complexity of these interaction mechanics for embolus transport in pulsatile arterial hemodynamics in multiple prior studies [36, 38]. The momentum response time is a key physical variable that determines this embolus-hemodynamics interaction mechanics. Momentum response time represents the time taken by a finite-sized inertial particle (*such as an embolus*) immersed in a fluid (*blood*) to respond to local changes in background fluid flow (*in this case, blood flow*). For this study, we considered thromboemboli in the diameter range of 0.5 mm - 4.0 mm, which leads to an estimated range of 0.004 sec - 0.24 sec for thromboembolus momentum response time (*refer to our prior work* [38]*, for instance, for further details*). Smaller emboli will have lower inertia and a smaller momentum response time, and can respond to and get entrained by the background blood flow faster. Conversely, larger emboli will have greater momentum response time, and will respond to hemodynamic forces slower, thereby deviating significantly from flow path. Consequently, embolus distribution under the LVAD driven jet flow presents with a strong size-dependency as observed from our analysis. This also explains the size-dependence in correlation coefficients with hemodynamic descriptors as noted in Section 3.6. We further note that this size-dependent dynamics; the complexity of the aortic flow with jet impingement, secondary circulation, vortex formation and helical flow; and the non-linear nature of the underlying hemodynamic forces on the emboli, lead to high variability in embolus distribution patterns and their departure from expectations based on flow distribution alone. This renders a major challenge to the understanding of embolic stroke or other cerebrovascular embolic events for patients on LVAD circulatory support. We have previously demonstrated the importance of understanding embolus-hemodynamics interactions to elucidate embolus distribution patterns across the Circle of Willis in the brain [37, 38]. The observations from this study further advance this argument for embolic events in patients on LVAD circulatory support.

### 4.3 Interplay of flow and graft anastomosis as a determinant for embolus destination

The direct comparison of thromboembolus distribution trends against variations in various hemodynamic descriptors, with varying anastomosis angles and LVAD pulse modulation, offers a novel perspective on hemodynamic underpinnings of stroke in LVAD circulatory support. For our investigation, the hemodynamic descriptors used are indicative of how the presence of LVAD alters hemodynamic patterns in the aorta compared to normal physiological flow scenarios [21], which is different than directly quantifying bulk descriptors of flow features under LVAD operation. When compared against thromboembolus distribution trends, we observed a strong negative correlation for wall shear stress descriptors. From the hemodynamics analysis presented in [21], the wall shear descriptor was highest for *Inc135* outflow graft anastomoses, where the graft inlet is directed away from the aortic valve. The LVAD outflow jet therefore is directed towards the distal aortic arch, and the jet impingement and post-impingement flow is less arrested compared to the other two *Inc* angle anastomoses. This increases the area over which the flow develops post-impingement, consequently increasing the average wall shear stress integrated over the aortic arch surface, as detailed in our prior work [21]. Concurrently, for the *Inc135* cases, as the emboli impinge on the distal aortic wall, they spread away from the point of impingement until being entrained in secondary vortices [57, 58]. This brings a significant fraction of emboli far downstream of the arch instead of traveling up the branching vessels into the cervical arteries towards the brain. This provides a hemodynamics based explanation for the strong negative correlation between altered state of wall shear stress due to LVAD flow, and thromboembolus movement towards the cervical vessels. We note that while this discussion addresses the connection between wall shear and embolus transport from the purview of jet impingement fluid flow in the aorta, wall shear stress by itself can have other mechanisms or roles to play in terms of pathological consequences (including disrupted endothelial mechanoregulation and altered endocrine signaling) in patients on LVAD support, which have not been discussed here. Additionally, while thromboembolus distribution towards the cervical vessels is lower for the *Inc135* anastomosis, a significant fraction of emboli travel distal to the impingement region down into the aortic root, leading to a higher accumulation of thromboemboli at the root driven by a larger stasis region (*as indicated in our prior studies* [21, 34]). This increased accumulation of emboli at the aortic root consequently increases the likelihood of thromboemboli traveling towards the cervical vessels in the event of intermittent aortic valve opening. These dynamic trends are clearly visible in the series of embolus trajectory animations provided in Supplementary Data. We further note that the effect of the anastomosis angle towards/away from the valve (that is, the *Inc* angle) has been extensively studied [22, 23], the effect of the anastomosis angle left/right of the heart (that is, the *Azi* angle) has been less explored. In our prior works, we presented detailed evidence on how the *Azi* angle influences aortic hemodynamic features. Here, we further advance that evidence to show how thromboembolus distribution is influenced by the *Azi* angle. We observe that the *AziNeg45* anastomosis generally presents high levels of thromboembolus distribution towards the cervical vessels, and for the *Inc45* and *Inc90* models, the embolus size dependence manifests differently compared to other *Azi* angles. One potential explanation for this is that for our model, the *AziNeg45* angle shows greater alignment with the centerline of the aortic arch, thereby better aligning the thromboemboli entering the aorta along the curvature of the arch, leading to distinct differences in thromboembolus distribution patterns. A visualization of this geometric alignment, as well as additional data visualization of thromboembolus distribution with respect to *Azi* angles are included in the Supplementary Data.

### 4.4 Clinical implications of findings

The data and insights presented here have multiple broad clinical implications. Thromboembolus source to destination propensity, and its ultimate distribution towards the brain, are direct determinants of stroke and other related cerebrovascular accidents. However, standard-of-care imaging and clinical work-up can not provide sufficient insights into embolus source-destination distribution, driven by embolus-hemodynamics interactions. The findings from this study provide valuable insights on the mechanisms governing embolic events, highlighting the underlying complex dynamic embolus-hemodynamics interactions. The observations indicate the importance of considering both: (a) the direct distribution of emboli entering the aorta towards the cervical vessels; and (b) the accumulation of emboli in the aortic root entrained in stagnated or slow moving flow - for characterizing embolus source-to-destination distribution under LVAD operation. We have quantified the thromboembolus transport as a function of LVAD outflow graft anastomosis, pulse modulation, embolus size, and source. Consequently, the underlying *in silico* framework can provide the basis for presurgical analysis of stroke risk post-implantation for individuals on LVAD support, as well as enable avenues for finding optimal patient-specific surgical configurations by analysing a number of virtual “candidate” surgical configurations and identifying ones with minimal predicted embolism risks. This will require further investigations leveraging the *in silico* methodology developed here, and remains an area of interest for future developments. The fact that thromboembolus movement towards the cervical vessels (and subsequently towards the brain) demonstrate a strong size-dependence is of additional clinical relevance. Specifically, smaller and larger emboli have different mechanisms of causing a stroke. Larger emboli get stuck in larger vessels and cause large vessel occlusion (LVO) in M2, A2 and P2 segments of the middle, anterior and posterior cerebral arteries. However, small emboli can perforate the distal beds causing lacunar infarcts [59, 60] and cerebral small vessel disease (CSVD) [61]. Our observations from the simulations presented here indicate that smaller thromboemboli entering the aorta from the LVAD outflow graft may have higher probability to transport towards the brain. Further systematic investigation comparing these predicted source-to-destination embolus transport patterns against noted stroke outcomes and stroke locations will be needed to further elucidate this across a larger cohort of patient cases.

### 4.5 Assumptions and limitations

Our *in silico* study was based on several key assumptions, with associated limitations, which are noted here. First, we assumed the embolus particle transport from aortic root as a source associated with continuous LO jet flow and did not consider the intermittent flow from AO jet from the aortic valve opening during ventricular systole. Additionally, we assumed that the vessel walls are rigid. Incorporating these additional structural features of intermittent valve opening and vessel wall deformations, within a parametric simulation study such as the one presented here, will be a computationally expensive endeavor and will require efficient numerical models for expanding such *in silico* approaches to resolve embolus-vessel-hemodynamics three-way interactions. This was not the scope of our study, but remains an area of active interest. However, the overall concept of mapping the source-destination relationship for emboli as outlined here, can be extended to these more resolved models as well. We remark here that, the approach of resolving vessel wall collisions using a Signed Distance Field, and the ability to virtually inject emboli as particles at a chosen source location (and over time) within the vascular model, are both key numerical modeling features presented here that can aid the transition into the aforementioned three-way interaction models. Second, we assumed a one-way coupling of the emboli particles with blood flow. This presumes that the blood flow influences the emboli, but the emboli themselves do not significantly alter the flow. This is a reasonable assumption considering that each embolus is modelled individually as a single particle released independently of the other, and that each individual embolus size is small in comparison to the vessel diameters they traverse. The alternative to this is a fully resolved computational model that numericall discretizes each embolus, which will be excessively expensive and prohibit the kind of Monte Carlo sampling study we have presented here. Third, our physics-based embolus-hemodynamics interaction model assumes that the emboli are spherical particles. This is an extensively-adopted modeling choice arising from the fact that presently there is no generalized physics-model for arbitrary non-spherical particle shapes. The derivation of a first-principles physics model of this nature will require significant additional theoretical and computational effort, which is beyond the scope of the study presented here. Moreover, we have not considered the embolus stretching, deformation, and deviation from sphericity governed by the flow-induced forces. Lastly, we have not considered here any modeling of turbulence, which can be of relevance at the impingement site of the LVAD Outflow jet. This remains another area of interest, which will require modifications of the physics-based model for embolus-hemodynamics interactions for turbulence, which have not been explored within the constraints and scope of this study.

## 5 Concluding remarks

In summary, we have presented an *in silico* framework for quantitative investigation of embolus transport, and thromboembolus source-destination mapping, towards characterizing stroke risks in patients on LVAD circulatory support. We conducted a systematic study with: (a) varying embolus sizes; (b) varying embolus release locations (sources); (c) varying LVAD outflow graft anastomoses; and (d) varying LVAD flow pulse modulation. The resulting simulations generated high resolution thromboembolus transport and distribution data, that was analyzed in terms of extent of thromboembolus distribution towards the cervical vessels supplying the brain, and extent of thromboembolus accumulation at the aortic root. We demonstrated size dependent distribution trends for thromboemboli as they travel towards the brain, and indicated how these distribution trends compare and correlate with purely hemodynamics-based descriptors such as wall shear, vorticity, and helicity. Such detailed insights can not be meaningfully obtained from existing imaging and clinical data alone, making this *in silico* approach highly valuable. Findings from our study also indicate how the *in silico* quantitative analysis provides insights that can enable pre-operative patient-specific planning to determine optimal LVAD graft anastomoses and pulse modulation. The established framework and analyses presented here can be further extended to now conduct larger patient cohort studies.

## Supporting information

Video-S1-smallest-size-combined

Video-S2-largest-size-combined

Video-S3-inc135aziNeg45low-size-combined

## Data Availability

All data produced in the present study are available upon reasonable request to the authors.

## Funding acknowledgements

This work was partly supported by a University of Colorado Anschutz-Boulder (AB) Nexus Research Collaboration Grant awarded to DM and JP; as well as partly supported by a National Institutes of Health Award (R21EB029736) awarded to DM. This work utilized resources from the University of Colorado Boulder Research Computing Group, which is supported by the National Science Foundation (awards ACI-1532235 and ACI-1532236), the University of Colorado Boulder, and Colorado State University.

## Author contributions

AS conducted all image-processing and hemodynamics modeling for the LVAD models. SM conducted all embolus transport simulations, including analysis and post-processing of all embolus distribution data. SM also drafted the manuscript. JP and EM guided study design, hemodynamic parameter selection, and clinical interpretation of thromboembolus transport data. DM designed the study, analysed and interpreted simulation data, and edited and finalized the manuscript in collaboration with SM, AS, EM, and JP. All authors reviewed and finalized the draft, and are in agreement regarding the final contents of the manuscript.

## References

1. Braunwald, E. Cardiovascular medicine at the turn of the millennium: triumphs, concerns, and opportunities. New England Journal of Medicine 337, 1360–1369 (1997).

2. Roger, V. L. Epidemiology of heart failure: a contemporary perspective. Circulation research 128, 1421–1434 (2021).

3. Savarese, G. & Lund, L. H. Global public health burden of heart failure. Cardiac failure review 3, 7 (2017).

4. Tsao, C. W. et al. Heart disease and stroke statistics—2023 update: a report from the American Heart Association. Circulation 147, e93–e621 (2023).

5. Rose, E. A. et al. Long-term use of a left ventricular assist device for end-stage heart failure. New England Journal of Medicine 345, 1435–1443 (2001).

6. Slaughter, M. S. et al. Advanced heart failure treated with continuous-flow left ventricular assist device. New England Journal of Medicine 361, 2241–2251 (2009).

7. Starling, R. C. et al. Results of the post-US Food and Drug Administration-approval study with a continuous flow left ventricular assist device as a bridge to heart transplantation: a prospective study using the INTERMACS (Interagency Registry for Mechanically Assisted Circulatory Support). Journal of the American College of Cardiology 57, 1890–1898 (2011).

8. Pagani, F. D. et al. Extended mechanical circulatory support with a continuous-flow rotary left ventricular assist device. Journal of the American College of Cardiology 54, 312–321 (2009).

9. Kirklin, J. K. et al. Seventh INTERMACS annual report: 15,000 patients and counting. The Journal of Heart and Lung Transplantation 34, 1495–1504 (2015).

10. Frontera, J. A. et al. Risk factors, mortality, and timing of ischemic and hemorrhagic stroke with left ventricular assist devices. The Journal of Heart and Lung Transplantation 36, 673–683 (2017).

11. Harvey, L. et al. Stroke after left ventricular assist device implantation: outcomes in the continuous-flow era. The Annals of thoracic surgery 100, 535–541 (2015).

12. Willey, J. Z. et al. Outcomes after stroke complicating left ventricular assist device. The Journal of Heart and Lung Transplantation 35, 1003–1009 (2016).

13. Acharya, D. et al. INTERMACS analysis of stroke during support with continuous-flow left ventricular assist devices: risk factors and outcomes. JACC: Heart Failure 5, 703–711 (2017).

14. Morgan, J. A. et al. Stroke while on long-term left ventricular assist device support: incidence, outcome, and predictors. ASAIO journal 60, 284–289 (2014).

15. Yuzefpolskaya, M. et al. The society of thoracic surgeons Intermacs 2022 annual report: focus on the 2018 heart transplant allocation system. The Annals of Thoracic Surgery 115, 311–327 (2023).

16. Plecash, A. R., Byrne, D., Flexman, A., Toma, M. & Field, T. S. Stroke in patients with left ventricular assist devices. Cerebrovascular Diseases 51, 3–13 (2022).

17. Koliopoulou, A., McKellar, S. H., Rondina, M. & Selzman, C. H. Bleeding and thrombosis in chronic VAD therapy: focus on platelets. Current opinion in cardiology 31, 299 (2016).

18. Eckman, P. M. & John, R. Bleeding and thrombosis in patients with continuous-flow ventricular assist devices. Circulation 125, 3038–3047 (2012).

19. Saltsman III, J. A., Ravin, R. A., Faries, P. L. & Tadros, R. Rapid progression of carotid artery atherosclerosis and stenosis in a patient with a ventricular assist device. Journal of Vascular Surgery Cases, Innovations and Techniques 2, 40–42 (2016).

20. Moulton, K. S., et al. PTEN deficiency promotes pathological vascular remodeling of human coronary arteries. JCI insight 3 (2018).

21. Sahni, A., McIntyre, E. E., Pal, J. D. & Mukherjee, D. Quantitative assessment of aortic hemodynamics for varying left ventricular assist device outflow graft angles and flow pulsation. Annals of Biomedical Engineering 51, 1–18 (2023).

22. Aliseda, A. et al. LVAD outflow graft angle and thrombosis risk. ASAIO journal (American Society for Artificial Internal Organs: 1992) 63, 14 (2017).

23. Callington, A., Long, Q., Mohite, P., Simon, A. & Mittal, T. K. Computational fluid dynamic study of hemodynamic effects on aortic root blood flow of systematically varied left ventricular assist device graft anastomosis design. The Journal of thoracic and cardiovascular surgery 150, 696–704 (2015).

24. Inci, G. & Sorgüven, E. Effect of LVAD outlet graft anastomosis angle on the aortic valve, wall, and flow. Asaio Journal 58, 373–381 (2012).

25. Karmonik, C. et al. Computational fluid dynamics in patients with continuous-flow left ventricular assist device support show hemodynamic alterations in the ascending aorta. The Journal of Thoracic and Cardiovascular Surgery 147, 1326–1333 (2014).

26. May-Newman, K., Hillen, B., Sironda, C. & Dembitsky, W. Effect of LVAD outflow conduit insertion angle on flow through the native aorta. Journal of medical engineering & technology 28, 105–109 (2004).

27. Prather, R., Divo, E., Kassab, A. & DeCampli, W. Computational fluid dynamics study of cerebral thromboembolism risk in ventricular assist device patients: Effects of pulsatility and thrombus origin. Journal of Biomechanical Engineering 143 (2021).

28. Chen, Z. et al. Shear stress and blood trauma under constant and pulse-modulated speed CF-VAD operations: CFD analysis of the HVAD. Medical & biological engineering & computing 57, 807–818 (2019).

29. Yang, N., Deutsch, S., Paterson, E. G. & Manning, K. B. Numerical study of blood flow at the end-to-side anastomosis of a left ventricular assist device for adult patients. Journal of biomechanical engineering 131, 111005 (2009).

30. Yang, N., Deutsch, S., Paterson, E. G. & Manning, K. B. Comparative study of continuous and pulsatile left ventricular assist devices on hemodynamics of a pediatric end-to-side anastomotic graft. Cardiovascular engineering and technology 1, 88–103 (2010).

31. Caruso, M. V. et al. A computational fluid dynamics comparison between different outflow graft anastomosis locations of Left Ventricular Assist Device (LVAD) in a patient-specific aortic model. International Journal for Numerical Methods in Biomedical Engineering 31, e02700 (2015).

32. Good, B. C., Deutsch, S. & Manning, K. B. Continuous and pulsatile pediatric ventricular assist device hemodynamics with a viscoelastic blood model. Cardiovascular engineering and technology 7, 23–43 (2016).

33. Mahr, C. et al. Intermittent aortic valve opening and risk of thrombosis in VAD patients. American Society for Artificial Internal Organs (ASAIO) Journal 63, 425 (2017).

34. Sahni, A., McIntyre, E. E., Cao, K., Pal, J. D. & Mukherjee, D. The relation between viscous energy dissipation and pulsation for aortic hemodynamics driven by a left ventricular assist device. Cardiovascular Engineering and Technology 14, 560–576 (2023).

35. Mukherjee, D., Jani, N. D., Selvaganesan, K., Weng, C. L. & Shadden, S. C. Computational Assessment Of The Relation Between Embolism Source And Embolus Distribution To The Circle Of Willis For Improved Understanding Of Stroke Etiology. Journal of Biomechanical Engineering 138, 081008 (2016).

36. Mukherjee, D. & Shadden, S. C. Inertial particle dynamics in large artery flows–Implications for modeling arterial embolisms. Journal of Biomechanics 52, 155–164 (2017).

37. Mukherjee, D., Jani, N. D., Narvid, J. & Shadden, S. C. The role of circle of Willis anatomy variations in cardio-embolic stroke: a patient-specific simulation based study. Annals of Biomedical Engineering 46, 1128–1145 (2018).

38. Roopnarinesingh, R., Leppert, M. & Mukherjee, D. Evidence and mechanisms for embolic stroke in contralateral hemispheres from carotid artery sources. Journal of the American Heart Association 12, e030792 (2023).

39. Vascular Model Repository https://www.vascularmodel.com/. Accessed: 2010-09-30.

40. Updegrove, A. et al. Simvascular: An Open Source Pipeline for Cardiovascular Simulation. Annals of Biomedical Engineering 45, 525–541 (2017).

41. Ku, D. N. et al. Blood flow in arteries. Annual review of fluid mechanics 29, 399–434 (1997).

42. Brooks, A. N. & Hughes, T. J. Streamline upwind/Petrov-Galerkin formulations for convection dominated flows with particular emphasis on the incompressible Navier-Stokes equations. Computer methods in applied mechanics and engineering 32, 199–259 (1982).

43. Franca, L. P., Frey, S. L. & Hughes, T. J. Stabilized finite element methods: I. Application to the advective-diffusive model. Computer Methods in Applied Mechanics and Engineering 95, 253–276 (1992).

44. Franca, L. P. & Frey, S. L. Stabilized finite element methods: II. The incompressible Navier-Stokes equations. Computer Methods in Applied Mechanics and Engineering 99, 209–233 (1992).

45. Taylor, C. A., Hughes, T. J. & Zarins, C. K. Finite element modeling of blood flow in arteries. Computer methods in applied mechanics and engineering 158, 155–196 (1998).

46. Ising, M. et al. Flow modulation algorithms for continuous flow left ventricular assist devices to increase vascular pulsatility: a computer simulation study. Cardiovascular Engineering and Technology 2, 90 (2011).

47. Maxey, M. R. & Riley, J. J. Equation of motion for a small rigid sphere in a nonuniform flow. The Physics of Fluids 26, 883–889 (1983).

48. Mei, R. An approximate expression for the shear lift force on a spherical particle at finite Reynolds number. International Journal of Multiphase Flow 18, 145–147 (1992).

49. Crowe, C., Schwarzkopf, J., Sommerfeld, M. & Tsuji, Y. Multiphase flows with droplets and particles. 2011. DOI 10, b11103 (2011).

50. Balu, A., Ghadai, S., Rauf Bingol, O. & Krishnamurthy, A. HyBoDT: Hybrid Bounded Distance Transforms of Trimmed NURBS Models. Journal of Computing and Information Science in Engineering 22, 041008 (2022).

51. Chueh, J. et al. Mechanical characterization of thromboemboli in acute ischemic stroke and laboratory embolus analogs. American Journal of Neuroradiology 32, 1237–1244 (2011).

52. Huang, C.-C., Chen, P.-Y. & Shih, C.-C. Estimating the viscoelastic modulus of a thrombus using an ultrasonic shear-wave approach. Medical physics 40, 042901 (2013).

53. Mukherjee, D., Jani, N. D., Narvid, J. & Shadden, S. C. The role of circle of Willis anatomy variations in cardio-embolic stroke: A patient-specific simulation based study. Annals of biomedical engineering 46, 1128–1145 (2018).

54. Carr, I. A., Nemoto, N., Schwartz, R. S. & Shadden, S. C. Size-dependent predilections of cardiogenic embolic transport. American Journal of Physiology-Heart and Circulatory Physiology 305, H732–H739 (2013).

55. John, R., Mantz, K., Eckman, P., Rose, A. & May-Newman, K. Aortic valve pathophysiology during left ventricular assist device support. The Journal of heart and lung transplantation 29, 1321–1329 (2010).

56. Da Rocha e Silva, J. G., et al. Influence of aortic valve opening in patients with aortic insufficiency after left ventricular assist device implantation. European Journal of Cardio-Thoracic Surgery 49, 784–787 (2016).

57. Liu, A. *Liquid jet impingement on a moving wall* PhD thesis (University of British Columbia, 2022).

58. Wang, W., Baayoun, A. & Khayat, R. E. A coherent composite approach for the continuous circular hydraulic jump and vortex structure. Journal of Fluid Mechanics 966, A15 (2023).

59. Arba, F. et al. Improving clinical detection of acute lacunar stroke: analysis from the IST-3. Stroke 51, 1411–1418 (2020).

60. Nakamori, M. et al. Lobar microbleeds are associated with cognitive impairment in patients with lacunar infarction. Scientific Reports 10, 16410 (2020).

61. Lee, W.-J. et al. Cerebral small vessel disease phenotype and 5-year mortality in asymptomatic middle-to-old aged individuals. Scientific reports 11, 23149 (2021).

