## Supplementary-Material-Information for "Understanding Embolus Transport And Source To Destination Mapping Of Thromboemboli In Hemodynamics Driven By Left Ventricular Assist Device"

### S1 Thromboembolus dynamics animations

We have included simulation animations, provided as an additional illustration of the simulation data to support discussions presented in the main manuscript. ***Video-S1-smallest-size-combined.mp4*** shows the thromboembolic particle transport dynamics for all 27 models. This animation illustrates thromboembolus trajectories for emboli entering from the LVAD outflow graft (*indicated in red*), and emboli originating at the aortic root (*indicated in blue*) for embolic particle diameter of 0.50mm. As discussed in detail in the main article, thromboembolus transport through LVAD outflow graft shows impingement in the inner aortic arch wall, in the initial phase of the animation. Likewise, ***Video-S2-largest-size-combined.mp4*** shows the thromboembolic particle transport dynamics for all 27 models. This animation illustrates thromboembolus trajectories entering from the LVAD outflow graft (*in red*), embmoli originating at the aortic root (*in blue*) for embolic particle diameter of 4.0 mm. These two animations, taken together, represent the smallest and largest thromboembolus sizes we have considered in our study, which further illustrates the aspect of size dependent thromboembolus transport, discussed extensively in the main manuscript. ***Video-S3-inc135aziNeg45low-size-combined.mp4*** shows the embolic particle transport dynamics for the *Inc135Azi-45* low modulation VAD flow model across all embolic particle sizes considered. This animation illustrates that thromboembolus trajectories shown in red for emboli entering from the LVAD outflow graft, and in blue for emboli originating at the aortic root in the *Inc135* model misses the cervical vessels as the jet directly impinges the distal part of the aortic arch. This further substantiates the observation of low extent of embolus transport towards the cervical vessels, as well as provides another illustration of the size dependent alteration of embolic trajectories.

### S2 Embolus size dependence as a function of azimuthal angles

We have already discussed in the main manuscript that embolus distribution is highly dependent on the variation of LVAD outflow graft anastomosis angles (*discussing primarily the outflow graft inclination angles towards/away from the aortic valve*). Here, we further present the embolus distribution dependence on outflow graft anastomosis in a different perspective in terms of the *Azi* angle, which remains sparingly investigated from a clinical perspective. Figure S1 illustrates the dependency of thromboembolus distribution fraction towards cervical vessels for varying embolus sizes. The panels are based on the different *Azi* angles considered for our study: *Azi0*, *Azi45* and *Azi-45*; for emboli entering from the outflow grasft as well as those originating at the aortic root. The mean distribution trend is represented using the green dashed line. The mean results show smaller emboli have a greater tendency to distribute towards the cervical vessels for all cases. However, for cases with *Azi-45*, we observe a few key differences. For *Inc45* and *Inc90* anastomoses, models with the *Azi-45* angle shows an initial increase in thromboembolus distribution towards cervical vessels based on emboluz size, which then decreases for larger particles. However, for *Inc135*, we observe that all anastomoses show a decrease in thromboembolus distribution towards cervical vessels with increasing embolus size. Likewise, Figure S2 illustrates the dependency of thromboembolus accumulation fraction in aortic root on embolus size. The panels are based on identical conditions as Figure S1. The results generally depict an opposite trend with Figure S1 as smaller emboli have a lesser tendency to be accumulated in the aortic root. Accumulation for *Inc45* is significantly smaller than *Inc135* for both sources which shows embolus impingement when graft angle is towards the valve, forces more embolus particles towards the cervical vessels and less accumulation in aortic root than when graft angle is away from the valve. The anastomoses with *Azi45* has lesser accumulation of embolus in the aortic root than *Azi0* and *Azi-45*.

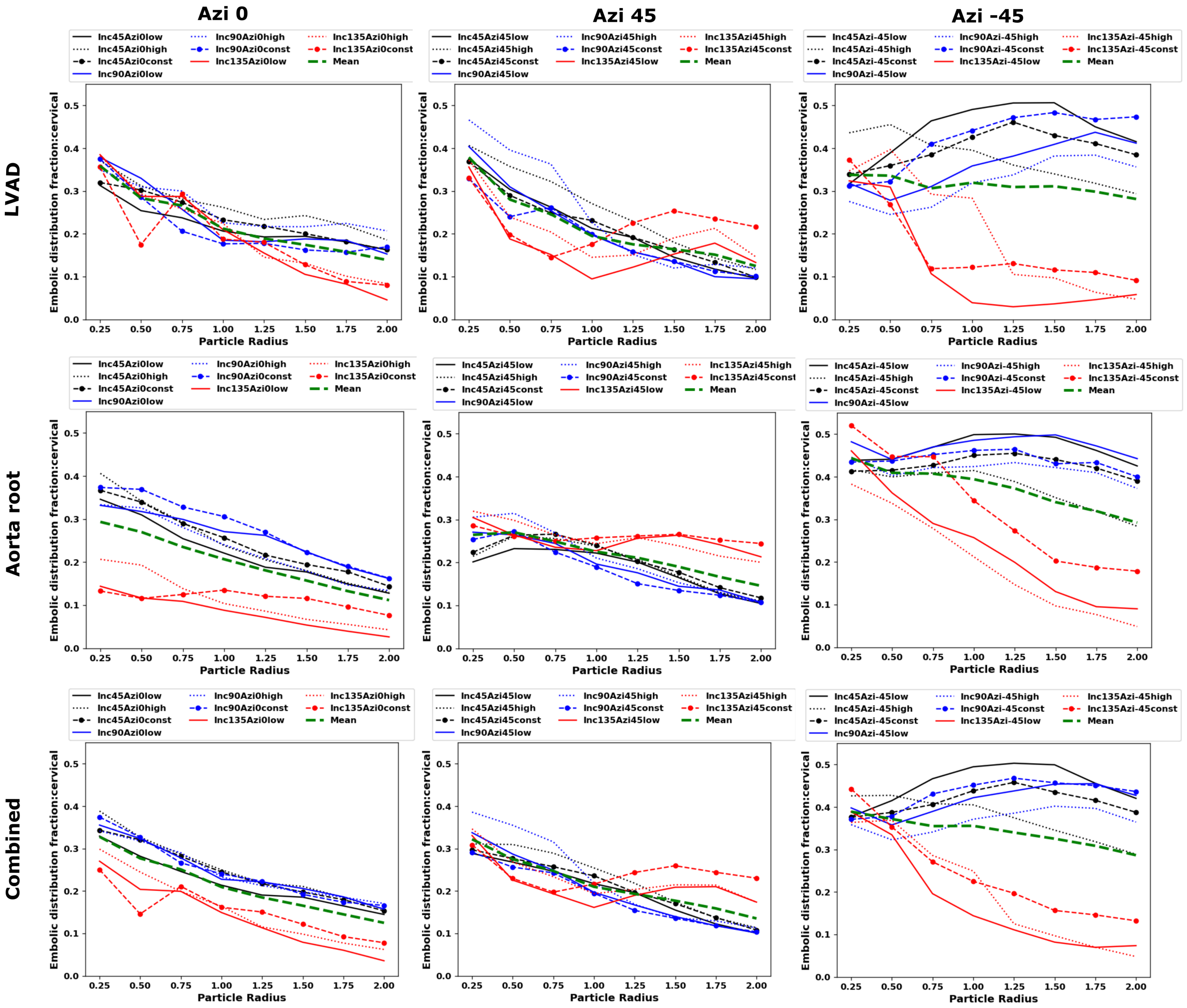


Figure S1: *Distribution of embolus particles along with embolus size based on Azimuthal angles for LVAD source, aortic root source and combined source towards cervical vessels. The color black, blue and red represents three different Inc angles: Inc45, Inc90 and Inc135 respectively. The solid line, dashed line and line with marker represents low pulse modulation, high pulse modulation and constant flow respectively. The mean of the plots in each panel is represented in dashed line with color green.*

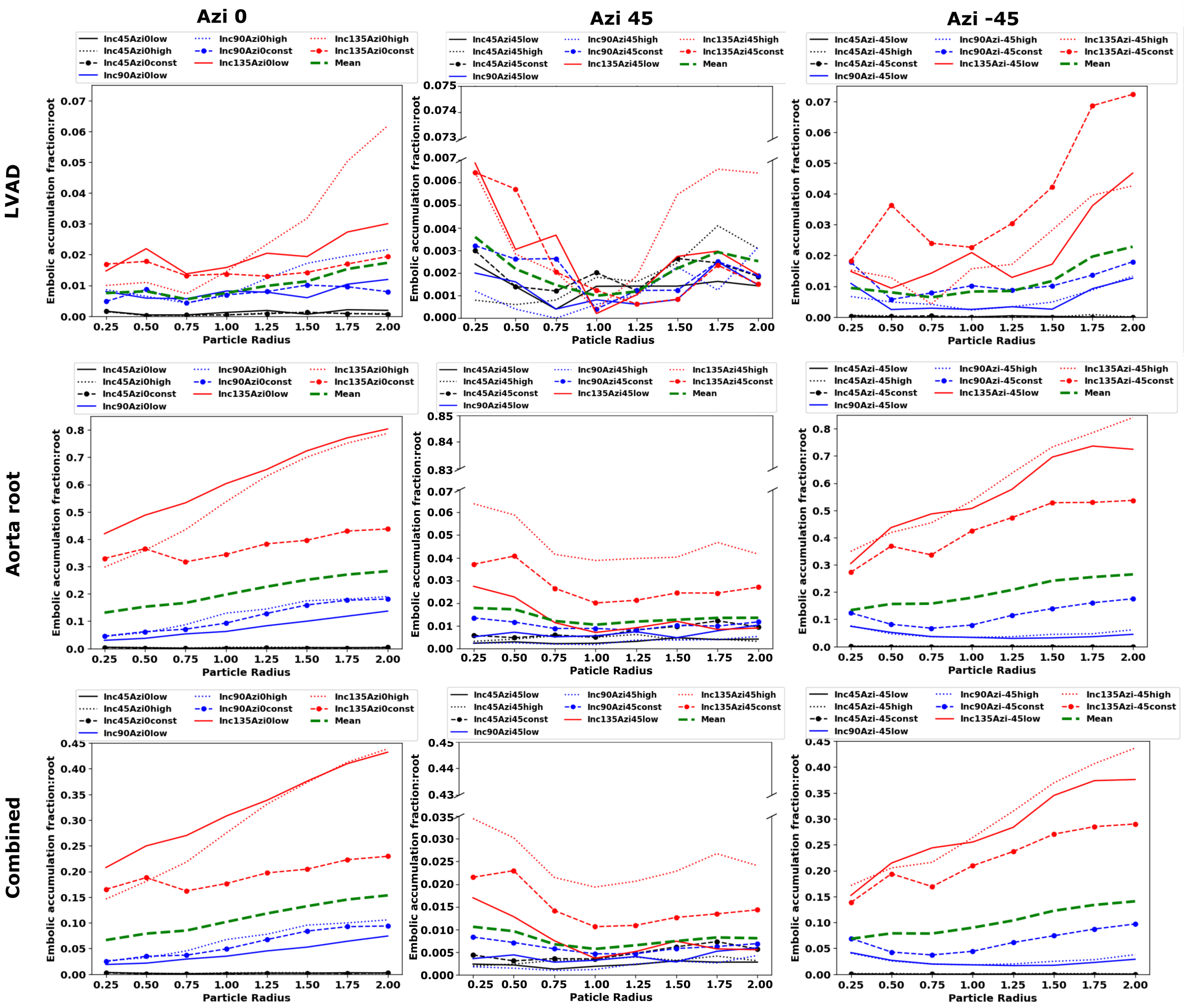


Figure S2: *Accumulation of embolus particles along with embolus size based on Azimuthal angles for LVAD source, aortic root source and combined source in aortic root. The color black, blue and red represents three different Inc angles: Inc45, Inc90 and Inc135 respectively. The solid line, dashed line and line with marker represents low pulse modulation, high pulse modulation and constant flow respectively. The mean of the plots in each panel is represented in dashed line with color green.*

### S3 Centerline projection for different azi angles

Figure S3 shows the comparison of centerlines generated for *Inc90* with different azimuthal angles, provided as a supplementary visualization to substantiate our discussion on interplay of graft anastomsis angle and flow for embolus transport as noted in Section 4.3 of the manuscript. The centerlines depict the geometric variations of LVAD attached to aortic arch, and we observe that LVAD outflow graft attached with an anastomosis angle of *Azi-45* is most aligned to the aortic arch curvature than *Azi0* and *Azi45*, potentially explaining therefore some of the specific thromboembolus dustribution trends observed from our simulation data for these anastomoses models.

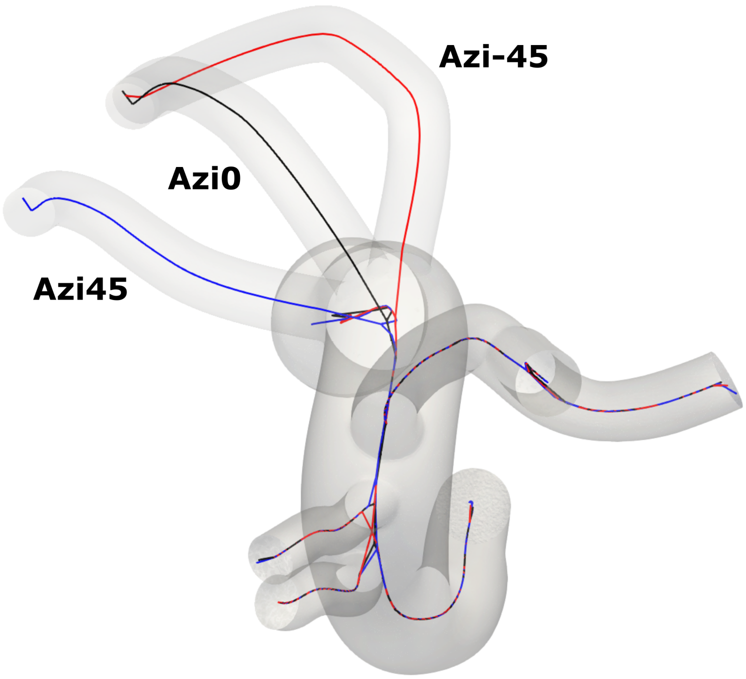


Figure S3: *Centerline projection (topview) for Inc90 case with three different Azi angles: Azi0, Azi45 and AziNeg45.*
